# A Cardiac Contouring Atlas of the Left Ventricle Myocardial Walls on CT

**DOI:** 10.64898/2026.05.06.26352374

**Authors:** Jielin Wei, Amir Abdollahi, Maximilian Knoll, Jennifer Furkel

## Abstract

**Background and purpose:** Precise manual annotation of the left ventricular myocardial (LVM) wall is essential for cardiac substructure research, wall-specific radiation dosimetry, and segmentation model development. However, existing radiotherapy-oriented atlases and conventional CT viewing planes lack an explicit framework for reproducible, wall-level LVM delineation. To address this gap, we developed an anatomy-guided manual segmentation protocol for delineating the five LVM walls on non-contrast-enhanced CT (NECT) or contrast-enhanced CT (CECT) scans.

**Materials and methods:** This protocol was developed using 60 chest CT scans from two prospective cohorts at Heidelberg University Hospital, including 50 CECTs from IMRT-MC2 cohort and 10 NECTs from MAGELLAN cohort. Manual contouring was performed in 3D Slicer. Segmentation rules were established through review by a radiation oncologist and a cardiology expert, based on the American Heart Association 17-segment model, and were tested on additional CT scans before final protocol definition.

**Results:** The protocol centers on three geometric steps: (1) defining the LV long axis using the endocardial apex and the center of the mitral annulus; (2) constructing an apical delimitation plane based on LV geometry; and (3) partitioning wall regions via intersections of the right ventricular and LV cavity centers in the short-axis view. This workflow enables structured segmentation of the anterior, septal, lateral, inferior, and apical LVM walls, supporting anatomically coherent 3D reconstruction.

**Conclusion:** This study provides contouring steps and a representative atlas as a methodological basis for standardized annotation, with potential applications in dose-mapping cardiotoxicity analysis and deep-learning modeling for radiotherapy.

## Introduction

The left ventricular myocardium (LVM) is not a homogeneous structure. Its regional organization is closely related to myocardial perfusion, ischemic vulnerability, infarct localization, and coronary arterial supply territories [1]. Thus, segmentation of the left ventricular (LV) into anatomically aligned wall regions has long been essential to cardiovascular imaging and cardiotoxicity analysis in radiotherapy[2–6]. The American Heart Association (AHA) 17-segment model [7] provides a widely used conceptual framework for LVM walls and has established a standardized link between myocardial regions and clinical interpretation. However, although the AHA model is highly valuable as a theoretical and clinical framework, it does not provide an operational contouring protocol for manual wall-level delineation on non-contrast-enhanced CT (NECT) or contrast-enhanced CT (CECT) scans.

The Oxford Cardiac Contouring Atlas for Radiotherapy [8] provides practical guidance for delineating cardiac substructures on treatment-planning CT. Several additional atlases have also been published, mainly focusing on contouring the whole heart and major cardiac cavities [9–11]. However, while these atlases are well suited for chamber-level and vessel-level segmentation, their direct application to subsegmenting left ventricular myocardium walls remains challenging. One difficulty is that the major anatomical axes of the heart mismatch with standard CT plane reconstructions: the wall anatomy of the left ventricular wall is best displayed in the short-axis orientation, whereas routine CT contouring is usually performed in the original axial view. This mismatch can introduce ambiguity in wall assignment, particularly at the transition from the basal and mid-ventricular myocardium to the apex. Moreover, although representative atlas slices may be available, they often do not provide sufficient detail for constructing a geometrically consistent 3D wall model across slices.

These limitations become particularly relevant when manual annotation is used as the basis for downstream AI model development. In that setting, the annotation protocol should not merely be visually plausible on individual slices. It should also provide a coherent geometric logic, allow consistent wall assignment, and support stable 3D wall reconstruction. Therefore, a protocol is needed that bridges three domains: the clinical interpretability of the AHA wall model, the practical requirements of CT-based manual contouring, and the structural consistency required for model training and 3D analysis.

To address this gap, we developed an anatomy-guided atlas for manual segmentation of the five left ventricular myocardial walls on CT, aiming to define a practical and anatomically coherent segmentation framework, including its conceptual rationale, anatomical definitions, and representative workflow, as a methodological basis for manual annotation and future standardization. We hypothesized that this protocol could support anatomically coherent wall-level delineation across CTs in both 2D and 3D views.

## Methods and Materials

A total of 60 chest CT scans were included from two prospective cohorts at Heidelberg University Hospital, comprising CECT scans from 50 patients in the IMRT-MC2 cohort and NECT scans from 10 patients in the MAGELLAN cohort. The IMRT-MC2 dataset includes CT scans from the multicenter phase III IMRT-MC2 trial (ClinicalTrial.gov NCT01322854), a prospective randomized clinical trial lead by Heidelberg University Hospital (2011-2015) including postoperative breast cancer patients for radiotherapy (25, 27-29). The MAGELLAN trial (NCT04925583), a prospective phase I study investigating MR-guided SBRT for ultracentral lung tumors at Heidelberg University Hospital (26). Cases with sufficient cardiac coverage and adequate image quality were included. Slice thickness ranged from 1.5 to 3 mm. Volumes of cardiac substructures were contoured using 3D Slicer (version 5.6.2, Slicer Solutions Inc., USA). A default window setting of width 350 and level 40 was adopted, with the option to adjust according to visualization and contouring needs. For the first set of 10 CTs in IMRT-MC2 cohort, the scans were reviewed by both radiation oncologist and cardiology expert to establish appropriate rules for segmenting the heart and substructures, based on the RTOG’s heart atlas [8] and AHA anatomical considerations [9]. After the segmentation rules were defined, a second set of 40 CTs was reviewed to test the feasibility and reproducibility of the segmentation rules. Subsequently, a third set of 10 scans was reviewed to assess whether these rules could also be applied to non-contrast-enhanced images. Following the predefined criteria of RTOG study, myocardial segmentation on NECT was performed using an average wall thickness of 1 cm [8, 12]. After iterative review across all 0 patients, the final segmentation definitions and the protocol were finalized and approved by the radiation oncologists and the cardiology expert.

## Results

The protocol consists of three major components. The first is construction of the LV geometric reference, centered on the definition of the long axis. The second is definition of the apical boundary, which separates the distal apical myocardium from the remaining wall structure. The third is wall-level partitioning in short-axis orientation, which assigns myocardium to the anterior, septal, lateral, inferior, and apical walls. After slice-wise annotation, the output is reviewed in 3D to verify overall continuity and anatomical plausibility. By this protocol, the LVM can be segmented into five wall-level components that are anatomically interpretable in both 2D and 3D rendering. We provide axial profile atlases of representative CECT scans, as well as 22 substructure segmentations of the heart, shown as Figure 1.

**Figure 1.**
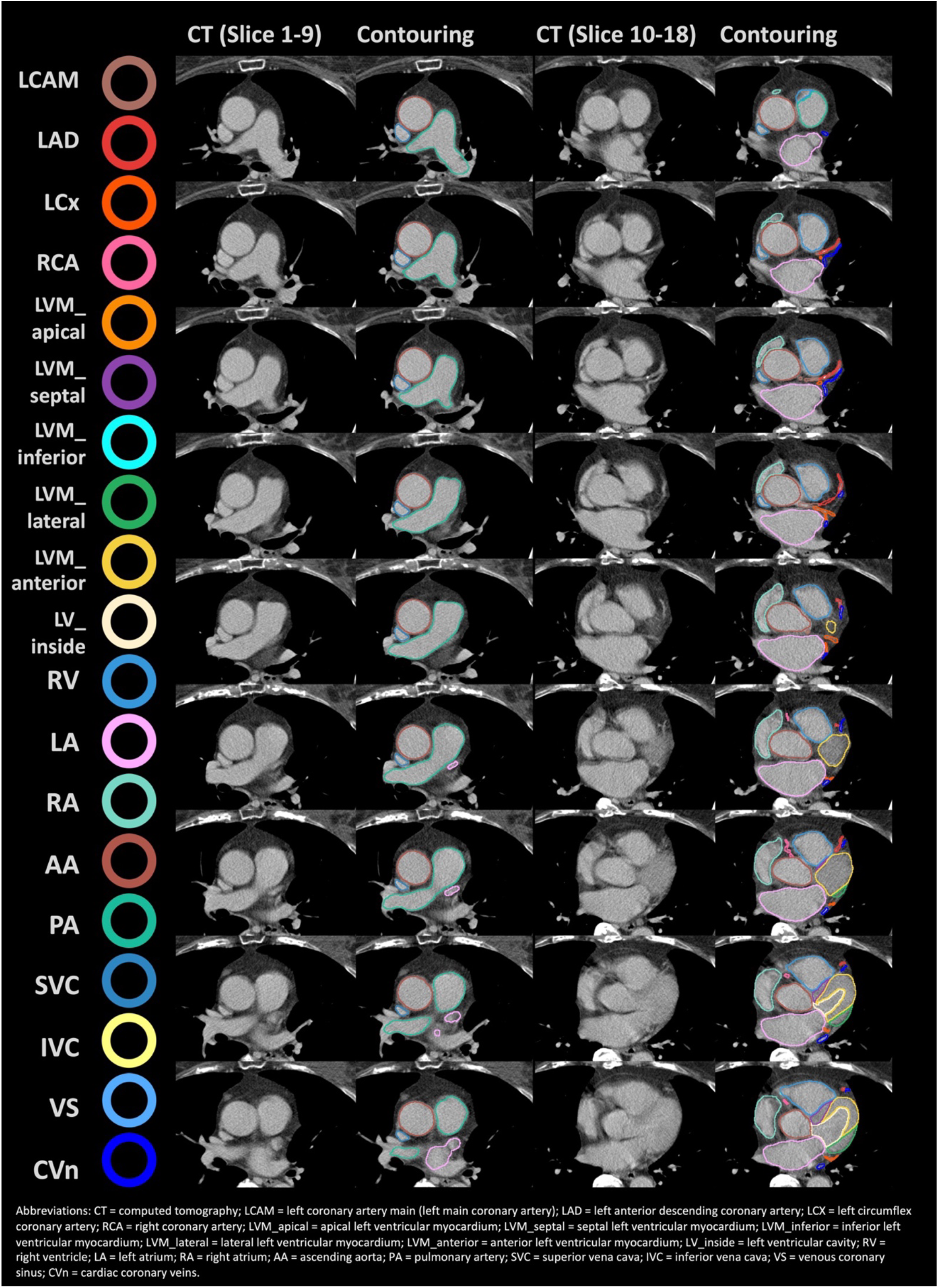
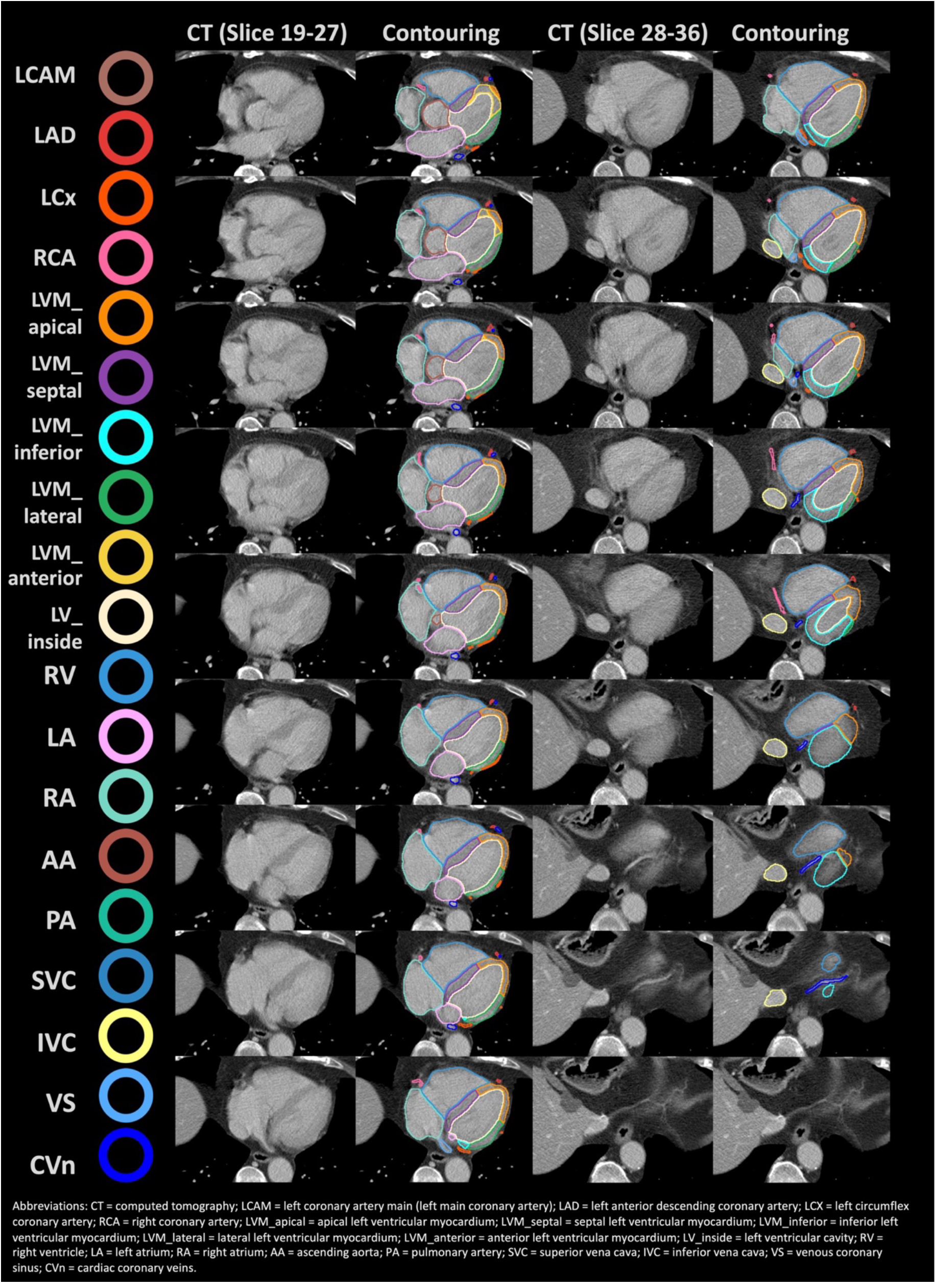
Axial contrast-enhanced CT images with manual contouring of the heart substructures (slice thickness 3mm).

### Definition of the LV Long Axis

The LV long axis serves as the primary geometric reference of the protocol. It is defined as the line connecting two chamber-oriented landmarks: the endocardial apex of the LV cavity and the center of the mitral annular region [13]. The starting point of the axis is the endocardial apex, not the outermost epicardial point [13]. The endpoint is the center of the mitral annular region, which represents the basal chamber reference rather than the outer contour of the left ventricle. Together, these two landmarks define the principal chamber direction axis of the LV.

### Construction of the Apical Delimitation Plane

After the LV long axis has been defined, an apical delimitation plane is constructed to separate the distal apical myocardium from the remaining ventricular wall. The plane is defined perpendicular to the LV long axis at the apical one-third point of the LV axis length. Myocardium distal to this plane is assigned to the apical wall, whereas myocardium proximal to the plane is subsequently partitioned into the septal, lateral, anterior, and inferior walls. By anchoring the apical boundary to a chamber-based geometric rule, the protocol provides an explicit rationale for separating the apical region from the remainder of the myocardium.

### Reorientation to LV-Oriented Short-Axis View

Following construction of the long-axis reference and apical plane, the image is reviewed in an LV-oriented short-axis view. This reformatted perspective is used because it more closely reflects the canonical arrangement of LV walls than the original axial view alone [14]. In this view, cross-sectional assessment of the myocardium becomes more anatomically intuitive.

### Partition of Non-Apical Myocardium into Four Wall Components

In the reformatted short-axis view, the non-apical myocardium is divided into four major wall regions: septal wall, lateral wall, anterior wall, and inferior wall. Following the 17-Segment AHA Myocardial Model [7, 13, 15, 16]., the LVM wall is divided into five anatomical regions based on the insertion points where the right ventricular (RV) and LV connect:

1. Anterior Wall (Segments 1 & 7)
2. Septal Wall (Segments 2, 3, 8, 9)
3. Lateral Wall (Segments 5, 6, 11, 12)
4. Inferior Wall (Segments 4, 10)
5. Apical Wall (Segments 13–17)

This partition is guided by the geometric relationship between the RV insertion points and the center of the LV cavity [13]. When connected with the LV cavity center, they provide a structural basis for dividing the myocardium into opposing sectors. On this basis, the myocardium can be partitioned into anatomically interpretable wall regions consistent with the canonical wall-level concept underlying the AHA myocardial model. The apical wall is treated separately, according to the apical plane defined above.

### Slice-Wise Contouring and 3D Review

Manual contouring is performed in a structured slice-wise manner. The operator contours slice by slice while examining the myocardium in the LV-oriented short-axis view. Each myocardial portion is then assigned to a predefined wall region following the established geometric framework. Annotation proceeds from the apex through the non-apical levels, maintaining wall assignment continuity across adjacent slices. The goal is to obtain both slice-level accuracy and 3D anatomical continuity across the ventricle. In practice, this demands iterative cross-checking between short-axis and 3D views, making slice-wise contouring part of a broader 3D workflow, not an isolated step. After contouring, the segmented wall regions are reviewed in 3D to ensure anatomical plausibility, regional continuity, and whole-structure coherence. This step is necessary because even individually acceptable slices can produce an irregular 3D structure.

## Discussion

This protocol was developed primarily to support manual reference annotation for model development, particularly for wall-level automated segmentation. In cardiology, it provides a structured framework for myocardial infarct localization by assigning infarcted regions to individual LV walls, thereby supporting regional cardiac injury analysis and future reproducibility research. Beyond that, in radiotherapy, it enables dose-mapping cardiotoxicity analysis for wall-specific dose assessment and heart-sparing planning.

Wall-level delineation of the left ventricular myocardium on CT remains methodologically challenging and limited. Although both the Oxford atlas proposed by Duane et al. [8] and the later Danish atlas by Milo et al. [11] represented important advances in radiotherapy-oriented cardiac contouring, both primarily provided conceptual or pictorial guidance rather than a fully operational geometric workflow for consistent wall assignment across slices. Duane et al. [8] established a five-wall LV concept, facilitating correlation between regional dose and clinically described sites of cardiac injury, whereas Milo et al. [11] simplified the left ventricle into four wall-related substructures (the apical excluded), emphasizing potential links to coronary blood supply and stenosis. This was an important step toward substructure-based cardiac dosimetry in radiotherapy. However, neither fully resolved the practical problem of how to assign wall boundaries consistently on CT slices or how to maintain anatomical coherence in 3D reconstruction.

In this protocol, we retained the five-wall framework of Duane et al. because it preserves a more stable geometric representation of the left ventricular wall, including a distinct apical wall. This distinction may be relevant in radiotherapy research, where regional dose distribution is often closely related to the 3D geometry rather than to a simplified vascular attribution alone. For example, in left-sided breast cancer radiotherapy with earlier 3D conformal tangential fields, the geometric position of LVM walls relative to heterogeneous dose deposition, with the apex at high risk. Furthermore, although the vascular orientation adopted by Milo et al. is clinically meaningful, a vessel-associated wall concept may be more difficult to standardize because coronary dominance patterns and branching anatomy vary substantially across individuals. By contrast, a geometry-based wall framework is a more readily transferable and more reproducible basis for wall-level contouring.

This work translates an existing conceptual model into an explicit contouring workflow. Specifically, the protocol defines the LV long axis from the endocardial apex to the mitral annulus center, derives an apical delimitation plane from LV geometry, and performs wall assignment in a rotated short-axis view using the relationship between the RV insertion points and the LV cavity center. This approach addresses a key limitation of prior atlases, namely the lack of procedural specificity regarding how wall-level assignment should be performed on a slice-by-slice basis.

This study has several limitations. First, the present work is primarily a methodological protocol based on retrospective study cohort analyses. It focuses on the conceptual framework and representative workflow, without quantitative interobserver evaluation, reproducibility, or external validation. Given the currently limited guidelines and reference standards for LV wall region delineation, quantitative benchmarking remains difficult. Additionally, manual annotation remains labor-intensive, and the protocol still requires anatomical familiarity and careful quality control. Nevertheless, the work provides an explicit, structured, and anatomically grounded workflow that can serve as a basis for future reproducibility studies, atlas refinement, and automated wall-level segmentation models.

## Conclusion

We describe an anatomy-guided protocol for manual segmentation of the five LVM myocardial walls on CT. The protocol is centered on a chamber-based LV long axis, a geometry-derived apical delimitation plane, and short-axis wall partitioning guided by the relationship between RV insertion points and the LV cavity center. By linking anatomical landmarks, geometric rules, and 3D structural review, this protocol provides a methodological basis for standardized heart wall-level annotation and future development of anatomically coherent automated segmentation models.

## Data Availability

All data produced in the present study are available upon reasonable request to the authors

## Author Contributions

Jielin Wei: Conceptualization, Data curation, Formal analysis, Funding acquisition, Investigation, Methodology, Project administration, Resources, Software, Validation, Visualization, Writing - original draft, Writing - review & editing. Jennifer Furkel: Supervision, Writing - review & editing, Conceptualization. Maximilian Knoll: Supervision, Writing - review & editing, Software. Amir Abdollahi: Supervision, Resources, Writing - review & editing.

## Acknowledgments

We thank investigators of IMRT-MC2 and MAGELLAN trials for data provision.

We thank Dr. Gordana Halec, Dr. Patrick Salome, Markus Ehle, David Kurz, Dr. Eva Meixner, Andreas Kudak, Matthias Dostal, Dr. Florian André, Dr. Mathias H. Konstandin, Dr. Lorenz Lehmann, Prof. Dr. Juliane Hörner-Rieber, Dr. Sebastian Regnery, and Prof. Dr. Dr. Jürgen Debus, for their support in facilitating access to the patient CT imaging scan used in this study and expert guidance in cardiac anatomy.

## References

[1] Heusch G, Libby P, Gersh B, Yellon D, Böhm M, Lopaschuk G, et al. Cardiovascular remodelling in coronary artery disease and heart failure. The Lancet. 2014;383:1933–43.

[2] Darby SC, Ewertz M, McGale P, Bennet AM, Blom-Goldman U, Bronnum D, et al. Risk of ischemic heart disease in women after radiotherapy for breast cancer. N Engl J Med. 2013;368:987–98.

[3] Lorenzen EL, Rehammar JC, Jensen MB, Ewertz M, Brink C. Radiation-induced risk of ischemic heart disease following breast cancer radiotherapy in Denmark, 1977-2005. Radiotherapy and Oncology. 2020;152:103–10.

[4] Gagliardi G, Constine LS, Moiseenko V, Correa C, Pierce LJ, Allen AM, et al. Radiation dose-volume effects in the heart. Int J Radiat Oncol Biol Phys. 2010;76:S77–85.

[5] van den Bogaard VA, Ta BD, van der Schaaf A, Bouma AB, Middag AM, Bantema-Joppe EJ, et al. Validation and Modification of a Prediction Model for Acute Cardiac Events in Patients With Breast Cancer Treated With Radiotherapy Based on Three-Dimensional Dose Distributions to Cardiac Substructures. J Clin Oncol. 2017;35:1171–8.

[6] Walls GM, O’Connor J, Harbinson M, Duane F, Mccann C, Mckavanagh P, et al. The Association of Incidental Radiation Dose to the Heart Base with Overall Survival and Cardiac Events after Curative-intent Radiotherapy for Non-small Cell Lung Cancer: Results from the NI-HEART Study. Clinical Oncology. 2024;36:119–27.

[7] Cerqueira M. Imaging. Standardized myocardial segmentation and nomenclature for tomographic imaging of the heart. A statement for healthcare professionals from the Cardiac Imaging Committee of the Council on Clinical Cardiology of the American Heart Association. Circulation. 2002:539.

[8] Duane F, Aznar MC, Bartlett F, Cutter DJ, Darby SC, Jagsi R, et al. A cardiac contouring atlas for radiotherapy. Radiother Oncol. 2017;122:416–22.

[9] Budoff MJ, Shinbane JS. Handbook of Cardiovascular CT: Springer; 2008.

[10] Feng M, Moran JM, Koelling T, Chughtai A, Chan JL, Freedman L, et al. Development and validation of a heart atlas to study cardiac exposure to radiation following treatment for breast cancer. International Journal of Radiation Oncology* Biology* Physics. 2011;79:10–8.

[11] Milo MLH, Offersen BV, Bechmann T, Diederichsen ACP, Hansen CR, Holtved E, et al. Delineation of whole heart and substructures in thoracic radiation therapy: National guidelines and contouring atlas by the Danish Multidisciplinary Cancer Groups. Radiotherapy and Oncology. 2020;150:121–7.

[12] Stolzmann P, Scheffel H, Leschka S, Schertler T, Frauenfelder T, Kaufmann PA, et al. Reference values for quantitative left ventricular and left atrial measurements in cardiac computed tomography. European radiology. 2008;18:1625–34.

[13] Selvadurai BSN, Puntmann VO, Bluemke DA, Ferrari VA, Friedrich MG, Kramer CM, et al. Definition of Left Ventricular Segments for Cardiac Magnetic Resonance Imaging. JACC Cardiovasc Imaging. 2018;11:926–8.

[14] Shalbaf A, Behnam H, Alizade-Sani Z, Shojaifard M. Automatic classification of left ventricular regional wall motion abnormalities in echocardiography images using nonrigid image registration. Journal of digital imaging. 2013;26:909–19.

[15] Yao B, Zhu R, Yang H. Characterizing the Location and Extent of Myocardial Infarctions With Inverse ECG Modeling and Spatiotemporal Regularization. IEEE J Biomed Health Inform. 2018;22:1445–55.

[16] Badano LP, Kolias TJ, Muraru D, Abraham TP, Aurigemma G, Edvardsen T, et al. Standardization of left atrial, right ventricular, and right atrial deformation imaging using two-dimensional speckle tracking echocardiography: a consensus document of the EACVI/ASE/Industry Task Force to standardize deformation imaging. Eur Heart J Cardiovasc Imaging. 2018;19:591–600.

